# Adverse events of colonoscopy in a colorectal cancer screening programme with faecal immunochemical test: a population-based community-based observational study

**DOI:** 10.1101/2020.05.09.20086389

**Authors:** Bernard Denis, Isabelle Gendre, Sarah Weber, Philippe Perrin

## Abstract

**Objectives:** Colonoscopy is considered a safe examination, serious complications being uncommon. Our study aimed to assess the adverse events of colonoscopy in a colorectal cancer screening programme with faecal immunochemical test (FIT) and to compare them with those of a previous programme with guaiac-based faecal occult blood test (gFOBT).

**Design:** Retrospective observational study

**Setting:** Population-based community-based colorectal cancer screening programme organised in Alsace, part of the national French programme, with FIT from 2015 to 2018 and gFOBT from 2003 to 2014.

**Participants:** All residents aged 50 – 74 years having a colonoscopy performed for a positive FOBT.

**Main outcome measures:** Adverse events were recorded through prospective voluntary reporting by community gastroenterologists and retrospective postal surveys addressed to persons screened.

**Results:** Of 9576 colonoscopies performed for a positive FIT, 6194 (64.7%) were therapeutic. Overall, 180 adverse events were recorded (18.8‰, 95% CI 16.1-21.5), 114 of them (11.9‰, 95% CI 9.7-14.1) requiring hospitalisation, 55 (5.7‰, 95% CI 4.2-7.3) hospitalisation >24hrs, and 8 (0.8‰, 95% CI 0.3-1.4) surgery. The main complications requiring hospitalisation were perforation (n=18, 1.9‰, 95% CI 1.0-2.7) and bleeding (n=31, 3.2‰, 95% CI 2.1-4.4). We observed one death (1 / 27,000 colonoscopies). Overall, 52 persons harbouring at least one adenoma ≥ 10 mm were detected for one adverse event requiring hospitalisation >24hrs. The rate of adverse events remained stable between gFOBT and FIT programmes.

**Conclusions:** The harms of colonoscopy in a colorectal cancer screening programme with FIT are more frequent than usually estimated, here six adverse events requiring hospitalisation >24hrs (three bleedings, two perforations), one necessitating surgery and 50 minor complications per 1000 colonoscopies. The price to be paid to save lives through colorectal cancer screening programmes is higher than what is stated in most pilots. Today, comparison between series dealing with colonoscopy-related adverse events is almost impossible.

## INTRODUCTION

Colorectal cancer (CRC) is the second most common cancer in Europe (500,000 estimated cases in 2018) and the second leading cause of death from malignant disease, resulting in 243,000 estimated deaths in 2018.^1^ Most are preventable. Several randomised controlled trials (RCTs) have demonstrated the efficacy of screening with guaiac-based faecal occult blood test (gFOBT) and flexible sigmoidoscopy on CRC mortality.^2^ Many countries have thus launched gFOBT CRC screening programmes.^3^ The efficacy of screening with a faecal immunochemical test (FIT) on CRC mortality is not demonstrated through RCTs but the superiority of FIT over gFOBT is now well established, so that many countries, including France, are now organising FIT CRC screening programmes instead of gFOBT programmes.^3^ However, the benefit-risk balance of these programmes is poorly evaluated.^4^ On one hand, none of the population-based screening programmes with gFOBT has been able to reproduce the CRC mortality reduction promised by the RCTs, and the expected benefit of population-based screening programmes with FIT has not been quantified yet. On the other hand, the risk of screening trials and programmes with gFOBT and FIT is poorly evaluated.^5^ We demonstrated that the adverse events (AEs) of colonoscopy were underestimated in all RCTs with gFOBT and are greater in a real-world programme.^6^ The rate of AEs requiring hospitalisation was estimated at 7.0% and 10.0% in two FIT CRC screening pilots, notably higher than the upper limit of 5‰ recommended by the European Society of Gastrointestinal Endoscopy (ESGE).^7–9^ Owing to the established risk factors (polypectomy, polyp size…) and since the neoplasia yield is significantly higher, AEs should be more prevalent in FIT-positive colonoscopies than in gFOBT-positive colonoscopies and, *a fortiori*, than in direct screening colonoscopies.^10,11^ Valid real-world data is needed to inform target populations, caregivers, policy-makers and funders.^4^

Our aim was to assess colonoscopy-related AEs in the CRC screening programme organised in France using an FIT and to compare them with those of the previous gFOBT programme.

## METHODS

We retrospectively analysed the AEs of all colonoscopies performed in residents undergoing colonoscopy for a positive FOBT within the population-based CRC screening programme organised in Alsace, a region in eastern France, 1.8 million inhabitants, part of the French national programme. The FOBT was a gFOBT from September 2003 to July 2014 and a quantitative FIT from May 2015 to January 2018.

### FOBT screening programme

A gFOBT CRC screening programme was initiated in Alsace in 2003. Its design has been previously described.^12,13^ Residents aged 50-74 years (0.57 million persons) were invited by mail every other year to participate. The gFOBT (Hemoccult II) was replaced by an FIT (OC-Sensor) in May 2015. The positivity threshold was set at 30 μg haemoglobin per gram (μg/g) faeces so that the positivity rate would be 4 to 5%. People with a positive FOBT were referred for colonoscopy.

All certified endoscopists participated in the programme. Colonoscopies were performed by community gastroenterologists under usual conditions, generally with sedation/anaesthesia provided by an anaesthetist. Diagnostic colonoscopy was defined as colonoscopy without intervention or with cold biopsy whereas therapeutic colonoscopy corresponded to any procedure with polypectomy, regardless of technique. Conventional adenomas, sessile serrated adenomas/polyps and traditional serrated adenomas were considered neoplastic polyps, whereas hyperplastic polyps were non-neoplastic polyps.

### Adverse events recording

AEs of all initial colonoscopies were recorded; those of surveillance colonoscopies were excluded. Information on AEs came from two main sources: prospective voluntary AE reports from gastroenterologists and seven retrospective postal surveys (every other year). For the postal surveys, all people who had undergone a colonoscopy received a mailed questionnaire with a pre-paid envelope for reply. The questionnaire asked for information on any colonoscopy-related AE and its consequences. The investigators reviewed all AEs with a phone call to the patient, and/or the general practitioner and/or the gastroenterologist. All colonoscopy reports and hospital charts concerning serious AEs were reviewed.

### Adverse event classification

AEs were classified according to the American Society for Gastrointestinal Endoscopy (ASGE) lexicon.^14^ All events definitely, probably and possibly related to colonoscopy occurring within 30 days of the colonoscopy were taken into account, whereas events unlikely related were not. An event that prevented completion of the planned procedure and/or resulted in admission to hospital, prolongation of an existing hospital stay, another procedure needing sedation/anaesthesia, or subsequent medical consultation was considered an AE. Unplanned events that did not interfere with completion of the planned procedure or change the plan of care were considered incidents. Bleeding that occurred and was managed during the procedure was considered as an incident. We added three indicators: AE requiring hospitalisation, AE requiring >24hrs hospitalisation, and AE necessitating surgery. The judgement on causality and severity was made by the two first authors (BD and IG).

### Ethical approval

Formal ethical approval was not required because it was considered a non-interventional study by the Est IV ethics committee (routinely collected data for the purposes of quality assurance within the screening programme).

### Statistical methods

We calculated the incidence of complications per 1000 colonoscopies and 95% confidence intervals (95% CI) using the binomial distribution (Wald). The Chi^2^ test was used to test for statistical significance by comparisons of proportions. All statistical tests were 2-sided. The significance threshold was set at 0.05. Statistical analysis was performed with Excel 2013 (Microsoft).

## RESULTS

### Colonoscopies and yield (Table 1)

Overall, 434,633 persons were invited, 194,975 (44.9%) participated and 9,450 (4.8%) had a positive FIT with subsequent 95.9% adherence to colonoscopy (9061 persons; mean age 63.0 years; SD 7.1). A total of 9576 colonoscopies were performed by 114 community gastroenterologists The most advanced lesion detected during therapeutic colonoscopies (64.7% of colonoscopies) was cancer (4.7%), neoplastic polyp (83.0%) and non-neoplastic polyp (12.3%). A total of 15,059 polyps were removed, that is a mean number of 2.4 polyps per therapeutic colonoscopy.

**Table 1:** Comparison between the two periods of screening with guaiac-based faecal occult blood test and quantitative faecal immunochemical test

|  | **gFOBT** | **FIT** |  |
| --- | --- | --- | --- |
|  | **(2003 – 2014)** | **(2015 – 2018)** | **p** |
| **Population** |  |  |  |
| Number of people | 17,152 | 9061 | - |
| Mean age (SD) years | 62.6 (7.0) | 63.0 (7.1) | <0.001 |
| Men n (%) | 9525 (55.5) | 5452 (60.2) | <0.001 |
| Postal survey participation n (%) | 11.519 (67.2) | 4605 (50.8) | <0.001 |
| **Colonoscopies** |  |  |  |
| Number of colonoscopies | 17,871 | 9576 | - |
| Therapeutic colonoscopies n (%) | 8700 (48.7) | 6194 (64.7) | <0.001 |
| Invasive cancer n (PPV) | 1086 (6.3) | 469 (5.2) | <0.001 |
| Advanced adenoma n (PPV) | 3957 (23.1) | 3209 (35.4) | <0.001 |
| Adenoma ≥ 10 mm n (PPV) | 3630 (21.2) | 2840 (31.3) | <0.001 |
| Adenoma n (PPV) | 6431 (37.5) | 5002 (55.2) | <0.001 |
| Non neoplastic polyps n (PPV) | 1637 (9.5) | 783 (8.6) | 0.02 |
| **Polyps** |  |  |  |
| Number of polyps | 18,443 | 15,553 | - |
| Number of polypectomies | 17,614 | 15,059 | - |
| Mean number of polypectomies per therapeutic colonoscopy (SD) | 2.0 (1.6) | 2.4 (2.0) | <0.001 |
| Proximal polyps n (% of polyps) | 5336 (28.9) | 5591 (35.9) | <0.001 |
| Polyps ≥ 2 cm n (% of polyps) | 1588 (8.6) | 864 (5.6) | < 0.001 |
| Surgery for benign polyps n (% per persons) | 247 (1.4) | 161 (1.8) | 0.04 |
| **Complications** |  |  |  |
| Incidents* n (‰) | 618 (34.6) | 349 (36.4) | 0.4 |
| Adverse events* n (‰) | 425 (23.8) | 180 (18.8) | 0.01 |
| Mild* AEs n (‰) | 342 (19.1) | 123 (12.8) | 0.001 |
| Moderate* AEs n (‰) | 57 (3.2) | 40 (4.2) | 0.2 |
| Severe* AEs n (‰) | 26 (4.5) | 17 (1.8) | 0.5 |
| AEs without hospitalisation and incidents n (‰) | 852 (47.7) | 415 (43.3) | 0.1 |
| AEs requiring hospitalisation n (‰) | 191 (10.7) | 114 (11.9) | 0.4 |
| AEs requiring hospitalisation >24hrs n (‰) | 94 (5.3) | 55 (5.7) | 0.6 |
| AEs necessitating surgery n (‰) | 19 (1.0) | 8 (0.8) | 0.6 |
| Perforations n (‰) | 20 (1.1) | 18 (1.9) | 0.1 |
| Bleeding n (‰) | 69 (3.9) | 43 (4.5) | 0.4 |

\* according to American Society for Gastrointestinal Endoscopy (ASGE) lexicon[15]
AE: adverse event; FIT: faecal immunochemical test; gFOBT: guaiac-based faecal occult blood test; PPV: positive predictive value; SD: standard deviation

### Adverse events

Overall, 180 (18.8‰ colonoscopies; 95% CI 16.1-21.5) AEs were recorded in 178 patients. The rate of AEs did not differ between men (18.4‰) and women (19.5‰) (p = 0.7) but increased significantly with age: 13.4‰ in people aged 50 to 59, 20.6‰ in those 60 to 69, and 23.5‰ in those 70 to 75 (p = 0.02). The rate of AEs requiring hospitalisation was 11.9‰ (95% CI 9.7-14.1), 16.6‰ for therapeutic and 3.3‰ for diagnostic colonoscopies (p<0.0001) (table 2). It was 5.7‰ (95% CI 4.2-7.3) for >24hrs hospitalisation, 7.3‰ for therapeutic and 3.0‰ for diagnostic colonoscopies (p=0.008). It varied from 0‰ to 50.0‰ depending on the endoscopist, with no correlation with the endoscopist’s adenoma detection rate or annual colonoscopy volume. The mean length of stay was 3.5 days (SD 5.5): overnight admission (n=59), stay of two or three days (n=23) and stay >3 days (n=32). The rate of AEs necessitating surgery was 0.8‰ (95% CI 0.3-1.4), 0.6‰ for therapeutic and 1.2‰ for diagnostic colonoscopies (p=0.4) (table 2). No death occurred.

**Table 2:**
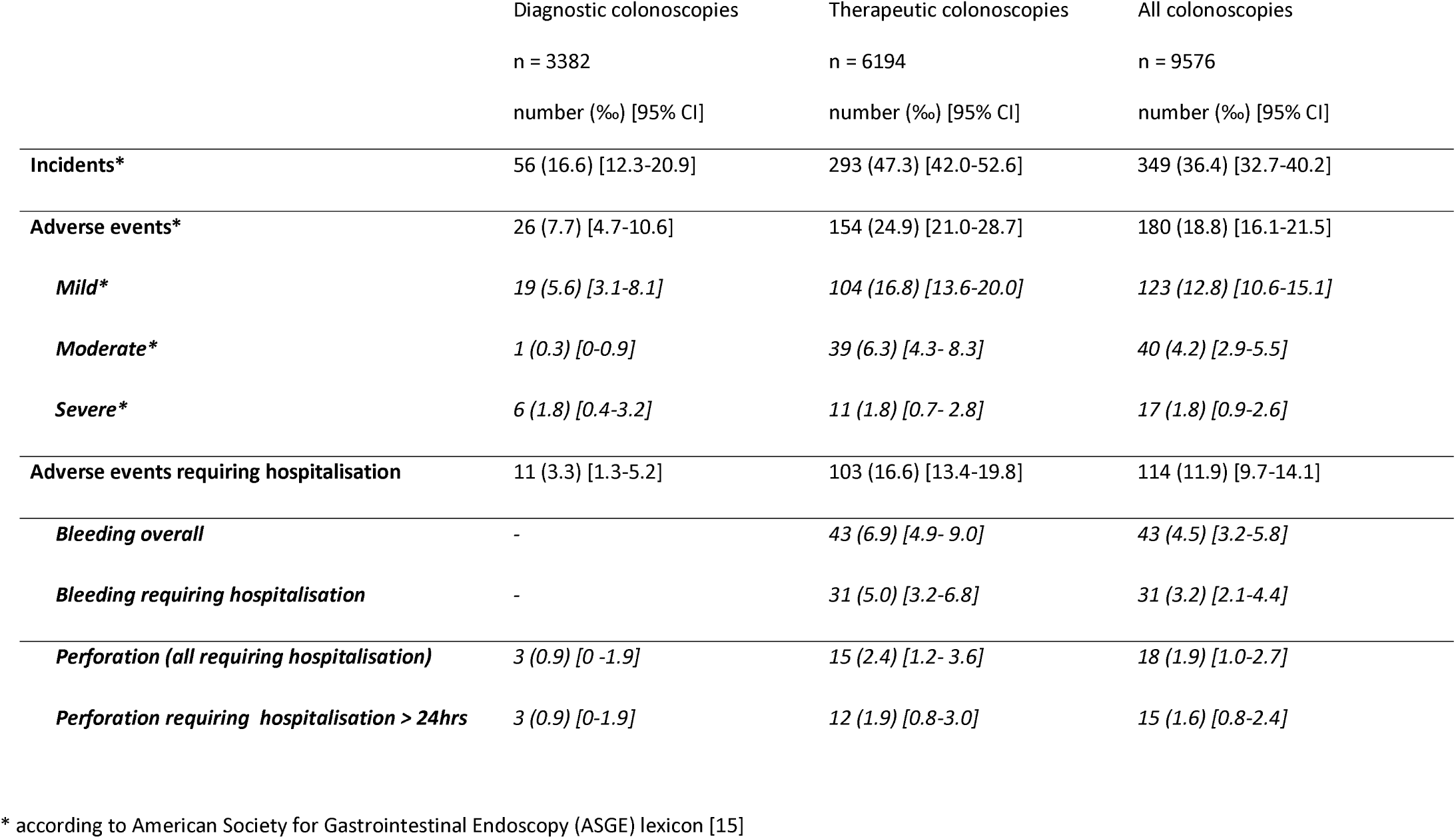
Classification of complications and their severity during the faecal immunochemical test period

One AE requiring >24hrs hospitalisation was encountered for 51.6 persons harbouring at least one adenoma ≥ 10 mm (58.3 one advanced adenoma).

### Perforations

A total of 18 perforations were recorded, that is a rate of 1.9‰ (95% CI 1.0-2.7), 3 (0.9‰) for diagnostic and 15 (2.4‰) for therapeutic colonoscopies (table 2). Severity was moderate in 5 cases (28%) and severe in 8 (44%). All required hospitalisation, 15 (1.6‰) hospitalisation >24hrs, and 7 (0.7‰) surgery (mean length of stay 12 days). Eleven (61%) perforations were diagnosed immediately; eight (44%) being immediately closed by the endoscopist (mean length of stay 2.5 days). The diagnosis of perforation was delayed in seven cases (39%) (mean length of stay 8.9 days), four of them being operated on. Thirteen (72%) perforations were caused by polypectomy, seven (39%) of them during a second colonoscopy performed by an “expert” for a large polyp (30 to 70mm) which a first endoscopist had been unable to remove. Three perforations were caused by mechanical disruption of the sigmoid colon wall due to the progression of the colonoscope and two perforations were caused by barotrauma. Four of them were operated on.

### Haemorrhages

Overall, 46 delayed haemorrhages were recorded, three were classified as an incident and 43 as an AE (4.5‰). Severity was moderate in 29 cases (63%) and severe in four (9%). All were caused by polypectomy, so that the rate of bleeding requiring hospitalisation was 3.2‰, 0‰ for diagnostic and 5.0‰ for therapeutic colonoscopies (2.3‰, 0‰ and 3.4‰ respectively for hospitalisation >24hrs) (mean length of stay 3.5 days) (table 2). Blood loss led to transfusion in nine cases (21%), repeat endoscopy in 30 (70%), endoscopic therapy in 23 (53%), and surgery in none (0%).

### Other adverse events (table 3)

Two splenic injuries were observed, one operated on and one treated by embolisation. One postpolypectomy syndrome was managed conservatively. Two cases of cardiac dysrhythmia secondary to hypokalaemia caused by the bowel preparation were encountered, as were one case each of myocardial infarction, pulmonary embolism, and aspiration pneumonia. Overall, 349 (36.4‰) incidents were recorded, so that the rate of incidents and AEs without hospitalisation was 43.3‰ (95% CI 39.3-47.4).

**Table 3:** Adverse events requiring hospitalisation during the two periods of screening with guaiacbased faecal occult blood test and quantitative faecal immunochemical test

|  | **gFOBT (2003 – 2014)** | **FIT (2015 – 2018)** | **p** |
| --- | --- | --- | --- |
|  | n = 17,871 colonoscopies | n = 9576 colonoscopies |  |
|  | number (‰) [95% CI] | number (‰) [95% CI] |  |
| **GASTROINTESTINAL AEs** | 175 (9.8) [8.3-11.2] | 104 (10.9) [8.8-12.9] | 0.4 |
| **Bleeding** | 54 (3.0) [2.2-3.8] | 31 (3.2) [2.1-4.4] | 0.8 |
| **Perforation** | 20 (1.1) [0.6-1.6] | 18 (1.9) [1.0-2.7] | 0.1 |
| **Post-polypectomy syndrome** | 7 (0.4) [0.1-0.7] | 4 (0.4) [0-0.8] | 0.9 |
| **Splenic injury** | 1 (0.1) [0-0.2] | 2 (0.2) [0-0.5] | 0.3 |
| **Surveillance** | 80 (4.5) [3.5-5.5] | 46 (4.8) [3.4-6.2] | 0.7 |
| **Other** | 13 (0.7) [0.3-1.1] | 3 (0.3) [0-0.7] | 0.2 |
| **NON-GASTROINTESTINAL AEs** | 16 (0.9) [0.5-1.3] | 10 (1.0) [0.4-1.7] | 0.7 |
| **TOTAL** | 191 (10.7) [9.2-12.2] | 114 (11.9) [9.7-14.1] | 0.4 |

AE: adverse event; FIT: faecal immunochemical test; gFOBT: guaiac-based faecal occult blood test

### Sources of information

A total of 4912 (51.3%) people answered the surveys. The pre-paid reply envelope was mistakenly omitted for the first FIT survey so that the answer rate was significantly reduced (33.5%) compared to that of the second FIT survey (70.0%) (p<0.00001). The rates of AEs, perforations and bleeding requiring hospitalisation >24hrs did not differ between the two surveys, whereas the rate of incidents and AEs without hospitalisation was significantly higher at the second survey (52.5‰ vs 34.6‰; p<0.01) (table 5). Overall, 85.1% of AEs requiring hospitalisation were notified by the gastroenterologists: 85.0% of perforations, 78.8% of bleedings, and 37.5% of non-gastrointestinal AEs.

**Table 4:**
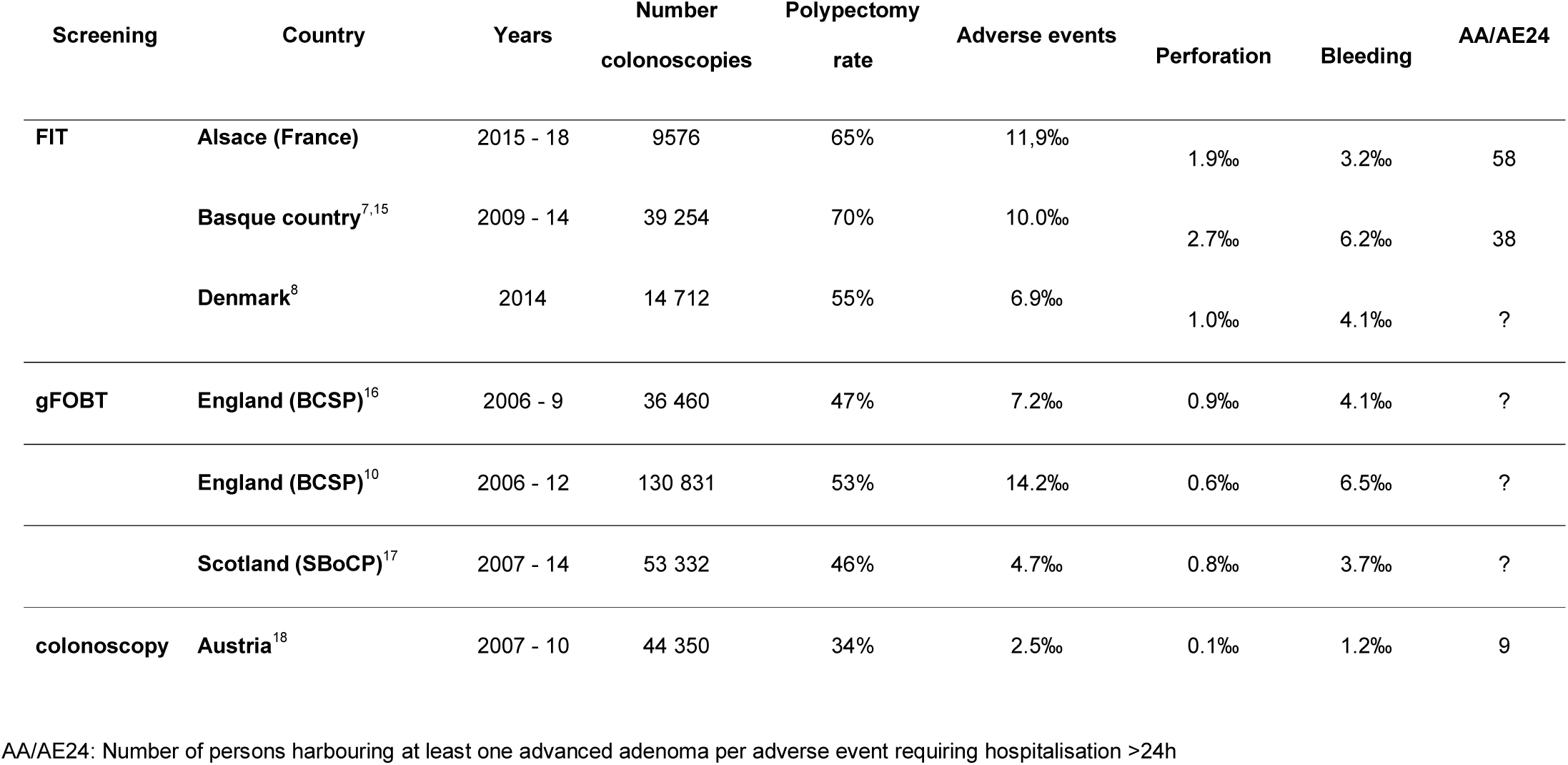
Adverse events requiring hospitalisation in different population-based studies of colorectal cancer screening programmes

| Screening | Country | Years | Number<br>colonoscopies | Polypectomy<br>rate | Adverse events | Perforation | Bleeding | AA/AE24 |
| --- | --- | --- | --- | --- | --- | --- | --- | --- |
| FIT | Alsace (France) | 2015 - 18 | 9576 | 65% | 11,9‰ | 1.9‰ | 3.2‰ | 58 |
|  | Basque country<sup>7,15</sup> | 2009 - 14 | 39 254 | 70% | 10.0‰ | 2.7‰ | 6.2‰ | 38 |
|  | Denmark<sup>8</sup> | 2014 | 14 712 | 55% | 6.9‰ | 1.0‰ | 4.1‰ | ? |
| gFOBT | England (BCSP)<sup>16</sup> | 2006 - 9 | 36 460 | 47% | 7.2‰ | 0.9‰ | 4.1‰ | ? |
|  | England (BCSP)<sup>10</sup> | 2006 - 12 | 130 831 | 53% | 14.2‰ | 0.6‰ | 6.5‰ | ? |
|  | Scotland (SBoCP)<sup>17</sup> | 2007 - 14 | 53 332 | 46% | 4.7‰ | 0.8‰ | 3.7‰ | ? |
| colonoscopy | Austria<sup>18</sup> | 2007 - 10 | 44 350 | 34% | 2.5‰ | 0.1‰ | 1.2‰ | 9 |

AA/AE24: Number of persons harbouring at least one advanced adenoma per adverse event requiring hospitalisation >24h

**Table 5:** Differences between the two surveys on screening with quantitative faecal immunochemical test

|  | **1st survey** | **2<sup>nd</sup> survey** | **p** |
| --- | --- | --- | --- |
| Number of colonoscopies | n = 4907 | n = 4669 | - |
| Response rate n (%) | 1646 (33.5%) | 3266 (70.0%) | < 0.01 |
| AEs without hospitalisation and incidents n (‰) | 170 (34.6) | 245 (52.5) | < 0.01 |
| AEs requiring hospitalisation n (‰) | 53 (10.8) | 61 (13.1) | 0.3 |
| AEs requiring hospitalisation >24hrs n (‰) | 23 (4.7) | 32 (6.9) | 0.2 |
| AEs necessitating surgery n (‰) | 3 (0.6) | 5 (1.1) | 0.4 |
| Perforations requiring hospitalisation >24hrs n (‰) | 6 (1.2) | 9 (1.9) | 0.4 |
| Bleeding requiring hospitalisation >24hrs n (‰) | 8 (1.6) | 14 (3.0) | 0.2 |
| Non-gastrointestinal AEs requiring hospitalisation >24hrs n (‰) | 4 (0.8) | 4 (0.9) | 0.9 |

AE: adverse event

### Comparison between gFOBT and FIT positive colonoscopies (table 1 and 3)

Several risk factors for complications increased significantly in the FIT period: rate of therapeutic colonoscopies, rate of proximal polyps and mean number of polyps per therapeutic colonoscopy (table 1). The overall rate of AEs decreased significantly from 23.8 to 18.8‰ (p = 0.01), whereas that of AEs requiring hospitalisation did not differ significantly (from 10.7 to 11.9‰, p = 0.4). The rates of AEs requiring hospitalisation related to therapeutic colonoscopies (from 19.8 to 16.6‰, p = 0.1), of gastrointestinal AEs (from 4.3 to 5.3‰, p = 0.2) and non-gastrointestinal AEs (from 0.4 to 0.6‰, p = 0.4) did not differ significantly. Endoscopic management of perforations increased significantly from 30% to 61% (p = 0.05) and all cases of bleeding were managed endoscopically in the FIT period.

One death occurred during the whole gFOBT and FIT period, that is one death for 27.447 colonoscopies (30-day mortality 3.6 / 100.000 colonoscopies) in a 57-year-old man.

## DISCUSSION

In our population-based community-based CRC screening programme with FIT, the rate of AEs requiring hospitalisation was 11.9‰ overall, 1.9‰ for perforation and 3.2‰ for bleeding. The rate of AEs requiring hospitalisation >24hrs was 5.7‰, 1.6‰ for perforation and 2.2‰ for bleeding. The rate of AEs necessitating surgery was 0.8‰, 0.7‰ for perforation and 0‰ for bleeding. Gastrointestinal AEs requiring hospitalisation and hospitalisation >24hrs remained stable during the gFOBT and FIT periods despite a significant increase in several risk factors for complications: the rate of therapeutic colonoscopies increased from 48.7% to 64.7%, the mean number of polypectomies per therapeutic colonoscopy from 2.0 to 2.4, and the rate of proximal polyps from 28.9% to 35.9%. Compared with the gFOBT period, the need for surgical management of AEs decreased significantly during the FIT period, from 70.0% to 38.9% for perforation and from 4.3% to 0% for bleeding. During the whole gFOBT and FIT period, we observed one death (30-day mortality 1 / 27,000 colonoscopies) and three splenic injuries (1 / 9000 colonoscopies).

The main strengths of our study are the population-based community-based setting along with the direct patient surveys. Indeed, this study is one of the first to assess the harms of colonoscopy in a population-based FIT CRC screening programme. It is also noteworthy that the details of all AEs were confirmed through direct access to hospital charts, colonoscopy reports, and, whenever necessary, phone calls to the gastroenterologist and/or the general practitioner and/or the patient. Albeit, this study is not without weaknesses. The response rate to the patients’ survey was 51.3%, low in comparison with the 83.7% of the NHS BCSP,^10^ and hospitalisation claims for the period could not be analysed, so that some AEs may have been missed, leading to a potential underestimation of the rate of AEs. However, the low response rate induced an underestimation of incidents and AEs without hospitalisation only, without consequence for AEs requiring hospitalisation, hospitalisation >24hrs and surgery. Therefore, underestimation of clinically relevant AEs is probably less of an issue. Another limitation is that the postal surveys were performed every other year, so that the delay between colonoscopy and survey could be long for some patients. Likewise, this delay may have induced an underestimation of incidents and AEs without hospitalisation as patients may have forgotten some minor complications.

The harm caused by colonoscopies in our FIT programme is in line with that recently reported by two other programmes (table 4).^7,8^ Their rates of 7.0‰ and 10.0‰, along with our 11.9‰ rate of complications requiring hospitalisation, are notably higher than the maximum standard of 5‰ for 7-day hospital admission/readmission rate recommended by the ESGE.^9^ They were higher than those usually reported, that is 0.5‰ for perforation and 2.6‰ for bleeding in a systematic review and meta-analysis of 21 studies along with those reported in another review of six studies on colonoscopies for stool-positive testing, i.e. 0.8‰ for perforation and 1.9‰ for bleeding.^19,20^ However, the comparison with such recommendations and aggregated figures is not pertinent as there is considerable heterogeneity in the literature. Today, it is almost impossible to compare different series dealing with colonoscopy-related AEs because indication and yield are different; AE nomenclature and sources of information are different; rates of referral for surgery are different. Even within a single programme, AE rates vary twofold: e.g. from 7.2‰^16^ to 14.2‰^10^ in the English Bowel Cancer Screening Programme (BCSP).

Neoplasia yield is correlated with the risk of complication and increases significantly depending on the indication for colonoscopy (symptoms, direct screening, positive gFOBT, positive FIT).^6,7^ The rate of therapeutic colonoscopies varies more than twofold between series, from 32.9% in the German colonoscopy screening programme^21^ to around 50% in gFOBT programmes and 55% to 70% in FIT programmes.^7,8^ A way to compare different series is to analyse separately diagnostic and therapeutic colonoscopies. For diagnostic colonoscopies, our perforation and bleeding rates requiring hospitalisation were 0.9‰ and 0‰, in line with those reported in the meta-analysis (0.4‰ and 0.6‰, respectively).^19^ For therapeutic colonoscopies, comparison is possible only between series having similar indications and neoplasia yields. Our 5.0‰ rate of bleeding requiring hospitalisation was significantly lower than the 9.8‰ reported in the meta-analysis.^19^ By contrast, our perforation rate was 2.4‰, significantly higher than the 0.8‰ in the meta-analysis.^19^ Our high rate is probably related to the high number of polyps per therapeutic colonoscopy (2.4), their large size (5.6% of polyps ≥ 2 cm), their proximal location (35.9%) and the low rate of surgical resection of benign polyps (1.8%). This level of neoplasia yield is seldom encountered, almost exclusively in FIT-positive colonoscopies. The rate of polyp(s) ≥ 2 cm was 2.1% in the English gFOBT BCSP.^22^ The rate of patients with at least one polyp ≥ 2 cm was 1.1% in the Austrian colonoscopy screening programme (7.4% in our programme).^18^

Colonoscopy techniques have evolved continuously towards more comfort, safety and ability to remove large lesions. Recent haemostasis and suture techniques allow endoscopic management of complications. Gastrointestinal AEs can be expected to be more prevalent today with AEs related to endoscopic mucosal resection of numerous, large and right-sided polyps. In fact, during our FIT period, all bleeding episodes were managed endoscopically. Moreover, the rate of perforations managed endoscopically increased significantly from 30% to 61% between the gFOBT and FIT periods. In the latter period, 39% of perforations were probably inevitable, caused by an “expert endoscopist” during large polypectomies. Almost half (45%) were diagnosed immediately and managed endoscopically, the median length of the hospital stay being 2.5 days. These figures indicate that in recent years in Alsace community gastroenterologists have improved their performance level significantly. One might wonder whether accredited gastroenterologists could perform better. In the French and Scottish screening programmes,^17^ colonoscopies are performed by any certified community gastroenterologist, whereas in England and the Netherlands they are performed by a set number of accredited gastroenterologists.

Our findings suggest that the rate of AEs is significantly higher in a community-based population-based FIT CRC screening programme than usually reported.^19,20,23^ The invited population should be informed for improved shared decision-making. Today, the leaflet accompanying the letter inviting the French population to participate in FIT CRC screening mentions in tiny characters that serious colonoscopy-related AEs are rare, estimated at 3‰. The word “serious” is not further explained. Maintaining this figure that underestimates by a factor of two the true prevalence of AEs is a kind of propaganda. It must be updated. We propose this statement “Colonoscopy-related adverse events, mainly bleeding and perforation, are rare. It is estimated that for 1000 colonoscopies performed for a positive FIT, six complications requiring hospitalisation longer than 24 hours and one complication necessitating surgery will occur”.

The ASGE lexicon is essential as it is almost the only one to propose a standardised nomenclature for AEs^14^ However, it is not satisfactory for common use:^6,10,16,23^ it is rather complex so that it has to be detailed every time it is used; it is best suited for the United States healthcare system; and certain criteria are somewhat arbitrary and open to criticism.^6^ Moreover, it does not define any specific method for identifying AEs. The less one seeks, the less one finds. All studies that rely on voluntary reporting underestimate actual complication rates.^8,24^ In our study, 15% of AEs requiring hospitalisation were not reported by gastroenterologists. In Germany, the AE rate was threefold higher in an audit than in the national colonoscopy screening registry.^24^ The Danish CRC screening database registered only 29.4% of recognised complications.^8^ A survey directed toward all patients should be mandatory for the publication of any article dealing with AEs. Moreover, should we still be calling a perforation occurring during a large polypectomy that is immediately diagnosed, closed and under surveillance < 48hrs in hospital a complication? There is an imperative and urgent need for a worldwide consensus on AE nomenclature, collection and reporting. An international task force should be created to formulate a series of recommendations, standardise definitions and categories, develop standardised methodology to search for AEs, and establish rules for reporting endoscopy-related AEs, for the same reasons and in the same way as were established the consensus on post-colonoscopy CRC and the STROBE statement.^25^ In anticipation, we adopted three simple, precise, self-explanatory indicators, all relevant for patient information: AEs requiring hospitalisation, as it is the most commonly used criterion to define clinically relevant complications; AEs requiring hospitalisation >24hrs, as we feel that an overnight admission for surveillance after colonoscopy (around 50% of our hospitalisations) is not a true complication; and AEs necessitating surgery.

Rate of referral for surgery must also be considered. An AE following the treatment of a large sessile polyp is attributed either to endoscopy or to surgery depending on the type of resection. Surgical resection-related morbidity-mortality is significantly higher than endoscopic polypectomy-related morbidity-mortality.^26,27^ Consequently, if the rate of referral for surgery for benign polyps is high, the individual endoscopist’s or CRC screening programme’s colonoscopy-related AE rate will be low while the overall AE rate will be high. Unfortunately, the rate of referral for surgery is never specified in articles concerning colonoscopy-related AEs and results of CRC screening programmes. Yet, 1) it is far from negligible as surgery for benign colorectal polyps represents 25% of surgeries for colorectal neoplasia in the USA,^28^ and 2) surgery-related morbidity-mortality is almost entirely preventable, provided the endoscopist who encounters a polyp he/she cannot manage personally refers the patient to an “expert endoscopist” rather than to a surgeon. “Expert endoscopists” are able to remove endoscopically more than 90% of large polyps.^29^ Overall, all studies concerning AEs of colonoscopy series and CRC screening programmes should mention their rate of referral for surgery for benign polyps (or, failing that, mention their surgery-related AEs). The rate of referral for surgery was 1.8% in our programme, about half that reported in Brittany (4.1%).^30^ Our low rate of surgical resection may partly explain our high rate of perforations following therapeutic colonoscopies.

The benefit-risk balance of CRC screening remains to be assessed correctly.^4,5^ At a time when most countries are organising CRC screening programmes, there is an urgent need for a worldwide consensus on standardised indicators. Overall, owing to the risk factors of colonoscopy-related AEs (polyp number and size) the risk is roughly proportionate to the benefit. We propose a new indicator that is the number of persons harbouring at least one adenoma ≥ 10 mm per AE requiring hospitalisation >24hrs (that could be abbreviated A10+/AE24). Between our gFOBT and FIT programmes, A10+/AE24 increased from 39 to 52. This indicator would be the cornerstone for assessment and comparison of the benefit-risk balance of different CRC screening programmes (e.g. between different countries) and strategies (e.g. between FIT and colonoscopy screening programmes). Unfortunately, this indicator cannot be calculated in any previous study, so we presented another close indicator in table 4, that is the number of persons harbouring an advanced adenoma per AE requiring hospitalisation >24hrs.

### Conclusion

AEs of FIT-positive colonoscopies are more frequent than usually reported, estimated in our programme at six AEs requiring hospitalisation >24hrs (three bleedings, two perforations), one necessitating surgery, and 50 minor complications per 1000 procedures. The invited population should be openly informed of these figures. Overall, there is a lack of transparency, both in the medical literature and in the information delivered to the population about the benefit-risk balance of CRC screening.^4^ Almost all the gastroenterology literature states that colonoscopy is safe, serious complications being uncommon.^19,20,23^ Overall, gastroenterologists have an incentive to maximise the benefit and minimise the risk of colonoscopy. Our results indicate that the price to be paid to save lives through CRC screening programmes is higher than what is stated in most pilots. Nevertheless, the benefit-risk balance of CRC screening remains favourable, estimated in our FIT programme at 52 persons harbouring at least one adenoma ≥ 10 mm per AE requiring hospitalisation >24hrs. And finally, the heterogeneity of series dealing with colonoscopy-related AEs is so enormous that comparison becomes almost impossible. There is an imperative and urgent need for a worldwide consensus on AE nomenclature, collection and reporting.

## Data Availability

All deidentified participant data are available upon reasonable request from IG

## Abbreviations

AE: adverse event
ASGE: American Society for Gastrointestinal Endoscopy
CRC: colorectal cancer
ESGE: European Society of Gastrointestinal Endoscopy
FIT: faecal immunochemical test
gFOBT: guaiac-based faecal occult blood test
NHS BCSP: National Health Service Bowel Cancer Screening Programme
RCT: randomised controlled trial

## Acknowledgments

The authors thank all the general practitioners who participated in this screening programme, the participating gastroenterologists and pathologists for their contributions and all the staff of ADECA Alsace (Association pour le dépistage du cancer colorectal en Alsace).

